# Seven-day COVID-19 quarantine may be too short: assessing post-quarantine transmission risk in four university cohorts

**DOI:** 10.1101/2021.05.12.21257117

**Authors:** Andrew Bo Liu, Dan Davidi, Hannah Emily Landsberg, Maria Francesconi, Judy T. Platt, Giang T. Nguyen, Sehyo Yune, Anastasia Deckard, Jamie Puglin, Steven B. Haase, Davidson H. Hamer, Michael Springer

## Abstract

**Background:** Despite rising rates of vaccination, quarantine remains critical to control SARS-CoV-2 transmission. COVID-19 quarantine length around the world varies in part due to the limited amount of empirical data.

**Objective:** To assess post-quarantine transmission risk for various quarantine lengths.

**Design:** Cohort study.

**Setting:** Four US universities, September 2020 to February 2021.

**Participants:** 3,641 students and staff were identified as close contacts to SARS-CoV-2-positive individuals. They entered strict or non-strict quarantine and were tested on average twice per week for SARS-CoV-2. Strict quarantine included designated housing with a private room, private bathroom and meal delivery. Non-strict quarantine potentially included interactions with household members.

**Measurements:** Dates of exposure and last negative and first positive tests during quarantine.

**Results:** Of the 418 quarantined individuals who eventually converted to positive, 11%, 4.2%, and 1.2% were negative and asymptomatic on days 7, 10 and 14, respectively. The US CDC recently shortened its quarantine guidance from 14 to 7 days based on estimates of 2.3-8.6% post-quarantine transmission risk at day 7, significantly below the 11% risk we report here. Notably, 6% of individuals tested positive after day 7 in strict quarantine, versus 14% in non-strict quarantine. Ongoing exposure during quarantine likely explains the higher rate of COVID-19 in non-strict quarantine.

**Limitations:** Quarantine should be longer for individuals using antigen testing, given antigen testing’s lower sensitivity than qPCR. Results apply in settings in which SAR-CoV-2 variants do not affect latent period.

**Conclusions:** To maintain the 5% transmission risk that the CDC used in its guidance, our data suggest that quarantine with qPCR testing 1 day before intended release should extend to 10 days for non-strict quarantine.

**Funding Source:** None.

## Introduction

Coronavirus disease 2019 (COVID-19), caused by SARS-CoV-2, has caused an unprecedented global public health crisis (1). Isolating infected individuals and identifying and quarantining their close contacts remain key strategies to mitigate the spread of SARS-CoV-2. Quarantine is used to separate individuals who might have been exposed to SARS-CoV-2 to minimize their risk of transmitting SARS-CoV-2 to other people. Despite rising vaccine distribution in some countries, the vast majority of the world remains unvaccinated. Thus, quarantine remains a critical part of the global response to control COVID-19 and will continue to be necessary for the foreseeable future.

Quarantine length is a balance: a short quarantine brings increased risk of transmission from individuals who are infectious after release, while a long one may increase transmission by reducing compliance, stretching public health systems, and imposing additional economic and psychological hardship (2). The US Centers for Disease Control and Prevention (CDC) initially recommended a 14 day quarantine period based on estimates of the upper bound of the SARS-CoV-2 incubation period (2,3). In November 2020, however, to account for the costs of long quarantine, CDC identified two shorter quarantine options as “acceptable alternatives” for asymptomatic individuals. The first is a 10-day quarantine period without testing. The second is a 7-day quarantine if a test done on days 5-7 is negative (2). Similarly, France instituted a 7-day quarantine and Belgium, Germany and Spain adopted 10 days (4); on the longer end, Chinese cities Beijing and Dalian adopted 21 days (5).

The second alternative guideline was derived from estimates that the exposed person would have about 16% (10-25%) and 5% (1-9%) risk of transmitting after days 5 and 7 post-exposure, respectively (2). These estimates were based on mathematical modelling of relative transmission risk over time, which was fitted to empirical observations of latent periods or simulated infectious periods from other models of within-host infection (6). For example, the first model predicts a 10-day infectious period based on a gamma density function (6). The few existing, evidence-based quarantine length guidelines are based on similar models of transmission risk (7) or on empirical estimates of incubation period (2). The uncertainty in transmission risk likely contributes to the lack of consensus regarding the optimal duration of COVID-19 quarantine (7).

Knowing more about the dynamics of SARS-CoV-2 infection can improve assessments of quarantine duration guidelines. If we know that the vast majority of people convert by RT-qPCR test before *n* days of exposure, individuals testing negative at day *n* are very unlikely to be SARS-CoV-2-positive afterwards. A previous study measured SARS-CoV2 positivity at different times in quarantined K-12 students; however, positive cases were few and students were tested infrequently (rarely before day 7), making it difficult to draw conclusions about the students’ first date of positivity and thus the transmission risk from 7-day quarantine (8).

Here we report the conversion times—the times between exposure and first becoming SARS-CoV-2-positive—for 418 university students and staff who were identified as close contacts, quarantined, and later tested positive between September 2020 and February 2021. Because SARS-CoV-2 conversion time closely correlates with latent period (9)—the time from exposure to infectiousness—these conversion times approximate the latent period, which informs transmission risk estimates.

The university quarantine conversion time data we collected provide empirical estimates of latent periods and transmission risk, testing previous models in real-world data. The four universities tested consistently and frequently, yielding high-resolution conversion time data. Additionally, many of these conversion times were measured for individuals in strictly enforced quarantine in isolation, in many cases with meal delivery, linens and self-care necessities provided. Therefore these conversion times come from individuals who largely complied with quarantine. Data from other quarantine settings might overestimate conversion time and incubation period due to additional exposures during the quarantine. Importantly, these data empirically test the mathematical models which initially informed the shortened 7-day quarantine guidelines.

## Methods

Four universities (Boston University, Duke University, Harvard University, and Northeastern University) reported data from 418 students and staff who were quarantined due to potential SARS-CoV-2 exposure and subsequently tested positive between September 2020 and February 2021. The data include presence of symptoms and dates of last exposure, last negative test during quarantine, first positive test during quarantine and symptom onset if symptoms occurred. These dates enable us to place the conversion date in the interval between the last negative and first positive test; we then compute the conversion time as the difference between the conversion date and exposure date.

University contact tracing protocols were based on CDC and local public health guidelines. Those who were identified as close contacts entered quarantine; if they lived on-campus, they entered “strict” quarantine, which included housing specifically designated for quarantine, with meal delivery, linens and self-care necessities provided. Testing was generally conducted twice per week, with minor variation in testing frequency between universities and between cases. The detailed testing, contact tracing, and quarantine protocols can be found in the Appendix.

To estimate a distribution from a set of uncertainty intervals of conversion times, we use a kernel density estimate (Figure 2). Each interval is transformed into a binomial (N, p=0.5) probability distribution kernel function centered at the interval midpoint, where N is the length of the interval in days. This binomial kernel was chosen because it has bounded support equal to the interval length, is symmetric and reflects our hypothesis that conversion times are more likely in the center of the interval than in the periphery. Results are similar if we use a uniform kernel (Appendix Figure 1). 95% confidence intervals were estimated based on Agresti-Coull and calculated by the R PropCIs package (10,11).

Our research protocol was approved by the IRB of the Harvard Faculty of Medicine (IRB20-0581), and we have data use agreements with Boston University, Duke University (DUA21-0149) and Northeastern University (DUA20-1481).

## Results

The conversion times for the 418 students and staff who entered quarantine after exposure and tested positive are shown in Figure 1. Each line segment shows the lower and upper bound of conversion time for one person, as measured from the last exposure date to the dates of the last negative (left point) and first positive tests (right point), respectively. If the last negative date was missing or occurred before the exposure date, the exposure date was taken to be the lower bound. Because SARS-CoV-2 conversion time closely correlates with latent period (9), these conversion times approximate the latent period as well. The universities’ frequent testing enabled us to pinpoint conversion time to within at most a 4-day interval in 78% of cases.

**Figure 1.**
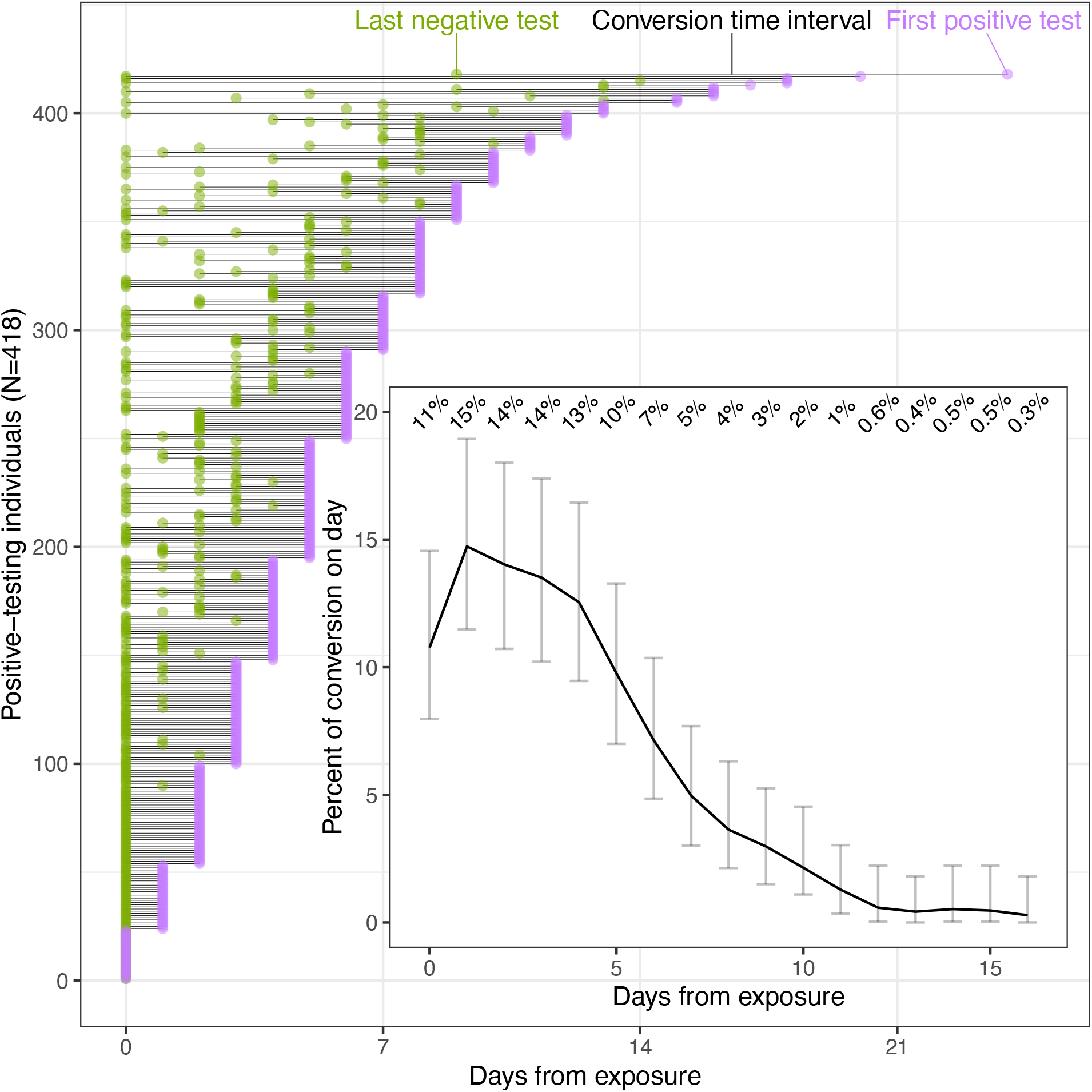
Determining conversion times of 418 university students and staff. Main figure: line segments for all 418 quarantined individuals show the lower (green dot) and upper bound (magenta dot) of conversion time based on the dates of the last negative and first positive tests, respectively. (If the last negative date was missing or earlier than the exposure date, the exposure date was used as the lower bound instead.) Inset is the probability distribution of conversion times, which highly correlate to latent periods. Error bars represent 95% confidence intervals (Methods). Numbers in the inset are the percent conversion on each day.

Nine percent of conversions happen after day 10 (Figure 1). There are significantly more conversions after day 10 in non-strict versus strict quarantine (11% versus 3%, respectively; p=0.01). Following up on several of these post-day 10 conversion cases revealed that, in many instances, the individual was actually re-exposed to a person with COVID-19 during their quarantine. This suggests that a higher rate of repeated exposures during quarantine can explain the longer conversion times seen in non-strict quarantine; thus strict quarantine data are more likely to provide unconfounded estimates of conversion times and latent periods in quarantine.

We then estimated the potential risk of conversion happening after release from quarantine by aggregating the above data (Figure 2). In the full dataset, 11% of positive-testing individuals were asymptomatic and had not yet converted on day 7 (95% CI 8.3-15%). Under the 7-day guideline, these 11% would have been released on day 7 and converted afterwards, reflecting post-quarantine conversion and therefore transmission risk. Similarly, 4.2% of converters were negative and asymptomatic on day 10 and 1.2% on day 14 (95% CIs 2.4-6.7%, and 0.35-3.1%, respectively).

Stricter quarantine was associated with shorter conversion times: in strict quarantine, 5.9%, 2.4% and <1% of individuals who converted did so after days 7, 10 and 14, respectively, compared to 14%, 4.9% and 1.7% in non-strict (95% CIs 2.5-12%, 0.14-7.3% and 0-4.3% for strict; 10-19%, 2.8-8.6% and 0.51-4.3% for non-strict).

**Figure 2.**
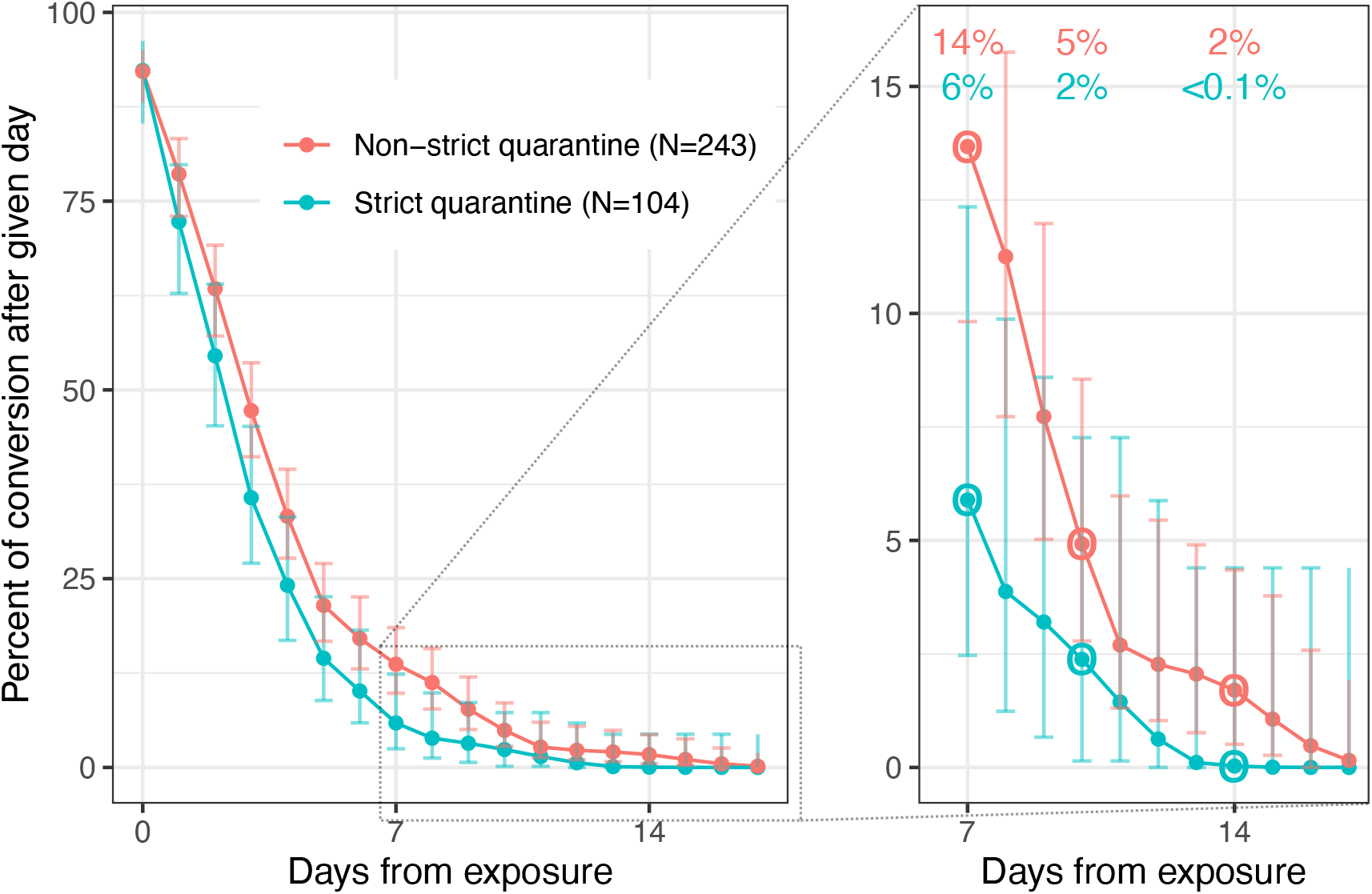
Transmission risk after release from test-based quarantines of various lengths for individuals in strict and non-strict quarantine. Individuals are released from quarantine if they receive a negative test and are asymptomatic. Complementary cumulative distribution plots show the chance of conversion for the 347 university students and staff for whom we had symptom data (out of 418 individuals); strict and non-strict quarantine are shown as blue and red, respectively. Values are shown for days 7 (corresponding to the CDC “quarantine with testing” alternative guideline), 10 (corresponding to the CDC “quarantine without testing” alternative guideline) and 14 (corresponding to the original CDC quarantine guideline). Strict quarantine included designated housing that consisted of a private room, private bathroom and meal delivery. Non-strict quarantine potentially included interactions with other household members. Error bars represent 95% confidence intervals (Methods). Inset shows transmission risks from day 7 onwards.

In Figure 2, we exclude the 71 people for whom we lack a last negative test date or symptom data. (Failing to exclude these 71 would have led to a distribution that is likely left of the true one, because we marked their conversion time lower bounds as 0 days after exposure.) Also, we assume no false negatives, which makes our transmission risk estimates conservative due to increased risk from releasing false negatives from quarantine.

We can also estimate transmission risks for quarantine based on antigen testing even though our conversion times are from RT-PCR data. We do so by assuming antigen testing would identify conversion 1 day later than a PCR test, which is reasonable given the lower sensitivity of antigen testing (even though the precise difference in sensitivity between the two tests varies by assay) (12). The details of the analysis, reasoning for the 1-day assumption and estimated transmission risks are shown in Appendix Figure 2 (analogous to Figure 2). As expected, the less sensitive antigen testing leads to a slightly higher post-quarantine transmission risk. For example, 14% of positive-testing individuals were negative and asymptomatic on day 7 with antigen testing, versus 11% with RT-qPCR.

All the above probabilities are conditional on individuals who converted to positive; probabilities conditional on exposure can be derived by multiplying by conversion rates. Of those who were exposed, 1.2, 0.5 and 0.1% converted asymptomatically after days 7, 10 and 14, respectively (95% CIs 0.91-1.7%, 0.26-0.74% and 0.039-0.34%). This is based on the 11% of exposed, quarantined individuals who converted (418 of 3,641; 95% CI 10.5-12.6%).

Similarly, of exposed individuals in strict quarantine, 10% converted (132 of 1,319), implying that 0.59, 0.24 and <0.001% of exposed individuals in strict quarantine converted after days 7, 10 and 14, respectively (95% CIs 0.25-1.2%, 0.014-0.73% and 0-0.43%). Of exposed individuals in non-strict quarantine, 12% converted (286 of 2,322), implying that 1.7, 0.59 and 0.20% of exposed individuals in non-strict quarantine did so after days 7, 10 and 14 (95% CIs 1.2-2.3%, 0.34-1.0% and 0.061-0.52%).

Thus, stricter quarantine was associated with a lower chance of testing positive (10% vs. 12%; p = 0.041 in 2-sided proportion test), supporting our hypothesis that non-strict quarantine contains individuals who were re-exposed during quarantine. This result, along with shorter conversion times in stricter quarantine, suggests that stricter quarantine more effectively reduces transmission.

## Discussion

Our results provide empirical evidence of the risk of transmission from people released from quarantine. In 418 quarantined, positive-testing university students and staff, 11% converted asymptomatically after day 7, 4.2% after day 7, and 1.2% after day 10. This means that 11% of transmission occurs after release from 7-day quarantine. 11% is substantially higher than the CDC’s mathematically modeled estimate of 2.3-8.6% (2). This comparison of latent periods has caveats: we have assumed 100% test sensitivity (underestimating true transmission risk); our data are biased towards young, high-socioeconomic status individuals in private universities; and conversion by RT-qPCR is tightly correlated with, but not the same as, infectiousness (9). With those caveats in mind, to limit post-quarantine transmission risk to 5% of total risk in a pre-B.1.1.7 university setting, our data suggest that quarantine with qPCR testing 1 day before intended release would need to extend to 10 days for non-strict quarantine and 7 days for strict quarantine. Most household or hotel quarantines probably more closely resemble our non-strict than strict quarantine (13,14). To achieve a post-quarantine transmission risk lower than 5% would require even longer quarantines (Figure 2).

Quarantine policies need to take into account many factors including socioeconomic cost, mental health, and the effectiveness of contact tracing. For example, the overall conversion rate in a given setting depends on the effectiveness of contact tracing and the stringency of guidelines for determining “close contacts.” In our dataset, 11% of quarantined individuals convert, meaning 1.2% of those who entered quarantine converted after day 7, 0.5% after day 10 and 0.1% after day 14; these percentages are likely very different in different settings. Hence, jurisdictions should consider our measurements of latent period as one factor among several in their overall quarantine guidance.

As noted in the US CDC guidance, quarantine is “intended to physically separate a person exposed to COVID-19 from others” (2). While both non-strict and strict quarantine intended to implement this isolation, strict quarantine was more effective. The chance of testing positive asymptomatically after day 7 was significantly lower in strict than non-strict quarantine, at 6% versus 14% (p = 0.06). Stricter quarantine also lowered the overall chance of testing positive from 12% to 10% (p = 0.041) and shortened conversion times such that in strict quarantine, 6%, 1% and <1% of individuals who converted did so after days 7, 10 and 14, respectively, compared to 16%, 5% and 1% in non-strict. Our study highlights that a substantial number of people are likely getting exposed during quarantine. A shorter quarantine with strict isolation may be as effective as a longer, more lax isolation.

The CDC 7-day quarantine guideline was created before at-home rapid antigen tests were available and before the known spread of SARS-CoV-2 variants like B.1.1.7. Early data suggest a longer duration of acute infection in those infected with B.1.1.7 versus non-B.1.1.7 SARS-CoV-2 (15). If conversion times for B.1.1.7 are also longer, then the quarantine durations suggested by this study’s results would be too short for those infected with this variant of concern. This could be solved by using genomic sequencing or variant-specific RT-PCR to differentiate B.1.1.7 from non-B.1.1.7 and advise longer quarantines for individuals infected with B.1.1.7, but this option may be expensive, and the additional turnaround time for sequencing may make such adaptive quarantine infeasible. Similarly, given the lower sensitivity of antigen testing, individuals who use antigen testing to test out of quarantine should quarantine for at least one additional day (16).

This study shows the importance of empirical validation of quarantine guidelines. As future variants emerge, similar analysis should be conducted to ensure the guidance remains relevant.

## Data Availability

Individual-level data not available due to privacy restrictions. Aggregated data available upon request.

## Acknowledgements

We would like to thank Becky Ward and Ron Milo for helpful discussions and feedback on the manuscript. We affirm that we have acknowledged everyone who contributed significantly to this study.

