## Supplement for "Seven-day COVID-19 quarantine may be too short: assessing post-quarantine transmission risk in four university cohorts"

### Appendix

#### University contact tracing, testing and quarantine protocols

Three of the four universities provided detailed testing, contact tracing and quarantine protocols; we reproduce those below.

##### Boston University

**SARS-CoV-2 testing.** Boston University developed four SARS-CoV-2 testing categories, related to risk of becoming infected on campus. Frequency of testing was based on these categories. These ranged from twice weekly for category 1 (on-campus undergraduates) to weekly testing for graduate students and faculty on campus to no testing for category 4 (e.g., students, faculty, or staff entirely off-campus).

**Contact Tracing.** The contact tracing protocol was based on CDC and Massachusetts Community Tracing Collaborative processes, with adaptation from BU academic programs and student input to capture the day-to-day intricacies of student life.

**Quarantine.** Students identified as a close contact (within 6 feet of an infected individual for a cumulative period of 15 minutes or more) had to quarantine for 14 days from the date of exposure. Based on updated guidance from the CDC, this period was reduced to 10 days of quarantine for asymptomatic individuals on November 20th. Isolation and quarantine rooms were pre-stocked with a 10 to 14 day supply of non-perishable foods and beverages, provided with linen and bedding, and basic self-care necessities a student would need while in quarantine or isolation. BU Dining Services delivered fresh meals and beverages four times a week. Undergraduate and graduate students in on campus quarantine are tested when they go into quarantine and have testing done twice a week. Sample collection supplies were delivered to their quarantine room for self-collection of anterior nares samples (via telehealth) and then the swabs were transported to the central campus testing center. In contrast, off campus students had to come into the symptomatic testing center twice per week to be tested. Anyone in quarantine had to have a negative test on or after day 8 (this began November 15th where prior to this quarantine extended for a total of 14 days) in order to be released on day 10.

##### Harvard University

**Quarantine, contact tracing and testing.** Undergraduates from Harvard College constituted the largest population of students living on campus during the fall semester. However, students were consistently housed in a single bedroom with a maximum of three people per bathroom. This allowed undergraduate students who needed to quarantine because of close contact to a positive Covid case (less than 6 feet for more than 15 minutes cumulative over 24 hours) to remain in their rooms during their quarantine. Graduate students living on campus had similar housing arrangements and were also able to quarantine in place. Meal deliveries occurred three times a day for this population through Dining Services. Students were also contacted on days 1, 2, 5, 8, and 10 (with an additional call on day 14 if necessary) by the contact tracing team and had contact with student support services periodically during their quarantine. Students in quarantine remained on a testing cadence with the guidance that quarantine could end at day 10 if tests were negative and students had no symptoms (per Massachusetts Department of Public Health guidelines). Students testing positive during quarantine were moved to isolation spaces and were required to complete the appropriate isolation timeline.

##### Northeastern University

**SARS-CoV-2 testing.** Northeastern University developed four SARS-CoV-2 testing cadences, related to risk of becoming infected and on-campus transmission.

1. Every three days:
   a. All students living in university housing
   b. All students who come to campus more than one day a week
   c. All undergraduate students living in off-campus residences in the neighborhoods surrounding the Boston campus.
2. Every four days:
   a. All staff and faculty who come to the Boston campus more than one day a week.
3. Any day when coming to campus:
   a. All staff, faculty, and students who come to the Boston campus one day a week or less.
4. Exempted:
   a. All staff, faculty, and students (except those included in #1) who work, teach, or learn completely remotely and do not come to the Boston campus.
   b. All staff, faculty, and students who have been confirmed with COVID-19 within the past 90 days.

*Staff includes vendor employees who work on the Boston campus.

**Contact Tracing.** The contact tracing protocol was based on the CDC and the Massachusetts Department of Public Health guidelines, under the supervision of the Boston Public Health Commission.

**Quarantine.** Students, faculty, and staff identified as close contacts (within 6 feet of an infected individual for a cumulative period of 15 minutes or more) had to quarantine for 14 days from the date of exposure regardless of test results until November 18th, 2020. On November 19th, 2020, Northeastern University changed the quarantine duration following the Massachusetts Department of Public Health guideline, to shorten the quarantine duration to 10 days if a PCR test taken on day 8 or later is negative and the individual has not experienced any symptoms. On December 9th, 2020, Northeastern changed the quarantine duration again following the updated Massachusetts Department of Public Health guideline based on the CDC guideline. Individuals can finish quarantine after 7 days if either PCR or antigen test taken on day 5 or later is negative and the individual has not experienced any symptoms. If the individual does not experience any symptoms, quarantine can end after 10 days without testing. If the individual has experienced any symptoms during the quarantine period, they have to quarantine for 14 days, regardless of test results.

Students who live in university housing and are identified as a close contact are required to move into Wellness Housing provided by Northeastern. Students who live in off-campus housing are strongly encouraged, but not required, to move into Wellness Housing. Students may choose to leave Wellness Housing before they finish quarantine if they have a place to safely finish quarantine and private means of transportation. This usually means their family home where they can drive to. The Wellness Housing room is not shared by multiple people. Each room for quarantine has a private bathroom only used by one person. All Wellness Housing rooms provide microfridge, bed linens, trash bags and a cleaning kit, toilet paper, soap, water, shelf-stable food, thermometer, pulse oximeter, and a blood pressure cuff. Northeastern's Dining Services deliver food and beverages once every day following an online order made by each student the day before. The Contact Tracing team places the order on behalf of the student on the first day.

**Testing During Quarantine.** Before November 18th, 2020, testing during quarantine was recommended to be taken on day 2, 5, 8, 11, 14 after the first date of exposure, and then again 14 days after the last date of exposure if it is different from the first date of exposure. An exposed individual's first date of exposure was determined to be the earliest day in the exposer's infectious period on which the index case and exposed individual had close contact. After November 18th, 2020, testing was recommended every three days during quarantine; earliest tests were allowed if they would permit an earlier end to quarantine. Testing during quarantine was provided at the symptomatic testing center on campus, and close contacts were allowed to go outside only when visiting the testing center or having medical emergencies. Close contacts quarantining far from the campus could get tested off campus.

#### Appendix Figures

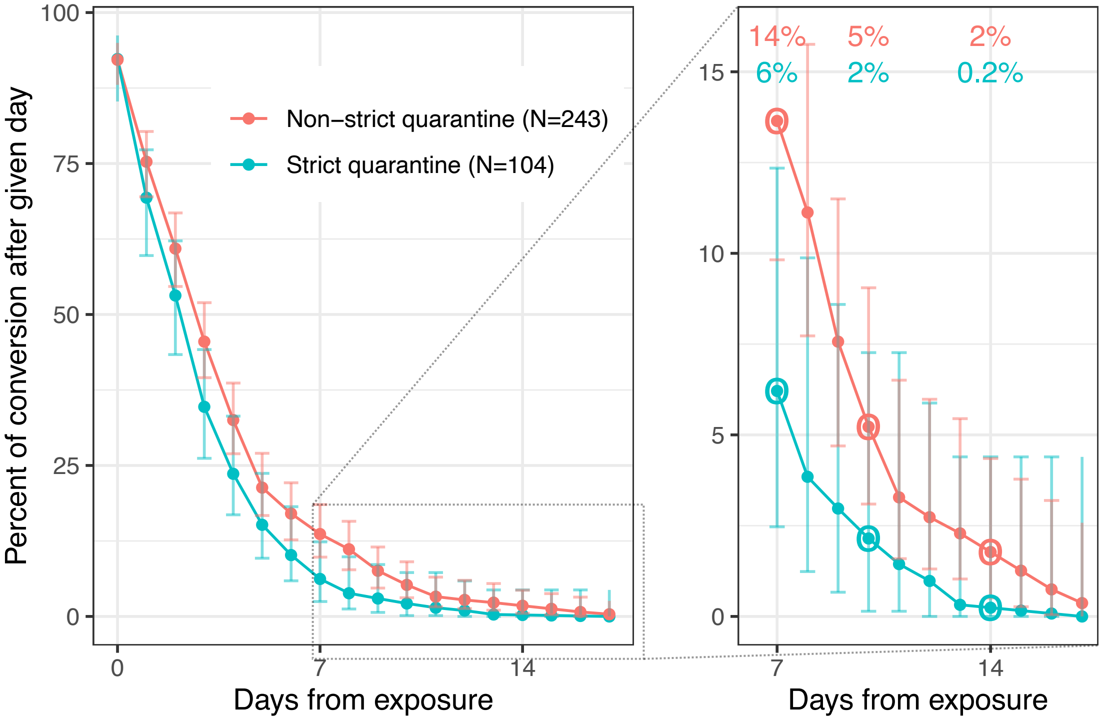


**Appendix Figure 1. Estimated transmission risk after release from test-based quarantines of various lengths for individuals in strict and non-strict quarantine.** This is analogous to Figure 2, except using a uniform kernel instead of a binomial kernel. The conversion rates are very similar to those computed from the binomial kernel.

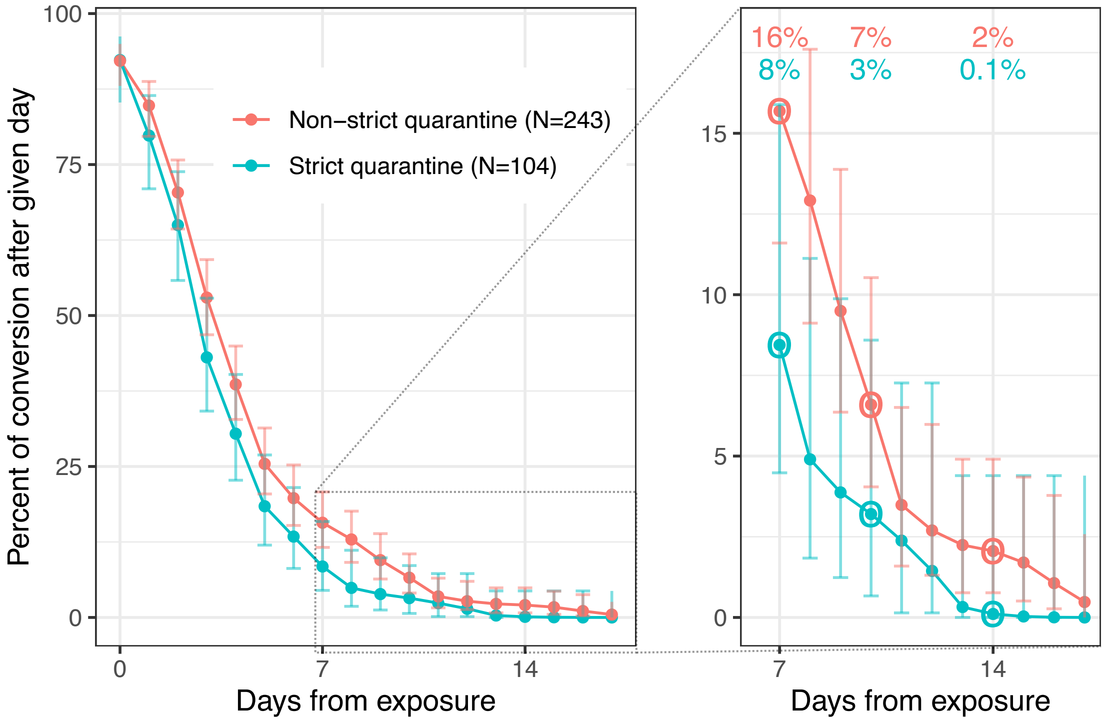


**Appendix Figure 2. Estimated transmission risk after release from antigen test-based quarantines of various lengths for individuals in strict and non-strict quarantine.** This is analogous to Figure 2, except an additional day of delay is added to each conversion time to model transmission risk from quarantine based on antigen testing. This one-day delay is based on the following assumptions/data: (1) SARS-CoV2 viral load rises by ~5 Ct per day in the proliferation stage of infection (16), (2) RT-qPCR is sensitive at 35 Ct, (3) antigen testing is nearly 100% sensitive at 25 Ct (12), and (4) RT-qPCR has a 1-day test turnaround while antigen testing has same-day turnaround. Thus someone who had 35 Ct on day *n* would test positive by RT-qPCR on day *n*+1 (due to 1-day turnaround) and positive by antigen testing on day *n*+2 (due to rise in 5 Ct per day for 2 days, with same-day turnaround).
